# Adaptive Post-Processing Recovers Most of the Gap to nnU-Net v2 in Head and Neck GTV Segmentation: A Paired Three-Arm HECKTOR 2025 Benchmark

**DOI:** 10.64898/2026.08.28.26361649

**Authors:** Ricardo Oyarzun Silva, Pablo Hernández Hernández

## Abstract

**Background:** Accurate delineation of the gross tumour volume (GTV) — primary tumour (GTVp) and nodal disease (GTVn) — on FDG-PET/CT is a critical step of head and neck radiotherapy planning. Comparisons between lightweight custom networks and the auto-configured nnU-Net v2 are usually reported as end-to-end pipelines, conflating the contribution of the network with that of the inference-time post-processing applied on top of it. We separated the two.

**Methods:** MiniUNet3D (custom 3D U-Net, 18.3 M parameters) and nnU-Net v2 (`3d_fullres`, 88.2 M parameters) were trained on the same 578 FDG-PET/CT cases (85/15 author-defined split of the HECKTOR 2025 Task 1 set, 8 centres) and evaluated on the same internal cohort. Three arms were compared pairwise: MiniUNet3D raw output at a fixed 0.5 threshold, MiniUNet3D with a locked adaptive post-processing pipeline, and nnU-Net v2. Comparisons used paired Wilcoxon tests with bootstrap confidence intervals, Bonferroni and Benjamini–Hochberg correction, and Cohen’s d; catastrophic failure (Dice <0.01) was compared with an exact McNemar test. Cases with an empty reference for a given target were excluded from that target’s analysis (n = 98 GTVp, n = 93 GTVn).

**Results:** With post-processing matched off, nnU-Net v2 was superior: median GTVp Dice 0.799 versus 0.592 (mean difference –0.244, 95 % CI –0.300 to –0.191; d = –0.88) and GTVn 0.774 versus 0.598 (d = –0.82). Post-processing raised MiniUNet3D to 0.800 (GTVp) and 0.738 (GTVn), recovering 79 % of that difference. Post-processed, MiniUNet3D matched nnU-Net v2 on GTVp Dice (p = 0.113) but remained inferior on nodal disease after Bonferroni correction (Dice p = 0.041; surface Dice p = 0.049). Catastrophic GTVp failures were 25/98 raw, 8/98 post-processed and 1/98 for nnU-Net v2 (McNemar p = 0.016). Inference took 34 s versus 78 s per case on the same GPU.

**Conclusions:** Post-processing recovered most, but not all, of the difference between the two models, and it did not confer robustness: an eight-fold higher rate of empty contours on small primaries persisted, which is the more consequential difference for planning safety. Pipeline comparisons reported without a post-processing ablation risk attributing to a network what post-processing supplied.

**Highlights:**

- **Three-arm paired benchmark** on HECKTOR 2025 Task 1: a lightweight custom 3D U-Net (**MiniUNet3D**, 18.3 M parameters) with and without adaptive post-processing, against the auto-configured **nnU-Net v2** (88 M parameters), on an identical training/validation split.
- **With post-processing matched off in both arms, nnU-Net v2 is decisively better**: median GTVp DSC 0.799 vs 0.592 (paired mean –0.244; Cohen’s d = –0.88; p < 0.001 after Bonferroni).
- **Adaptive post-processing recovers most of that gap**: it adds **+0.192 DSC** to MiniUNet3D on GTVp, **79 %** of the 0.244 model-level difference. Post-processing is thus slightly smaller than the model gap itself, but three to four times larger than what remains of it afterwards (0.051 – 0.062).
- **The gap is not fully closed.** Even post-processed, nnU-Net v2 retains a small but Bonferroni-significant advantage on nodal disease (GTVn DSC p = 0.041; sDSC@2mm p = 0.049) and an **eight-fold lower catastrophic failure rate** (1/98 vs 8/98; McNemar exact p = 0.016).
- Practical implication: a team already committed to a lightweight custom network can **recover most of what that choice costs at the inference stage, without retraining** — but post-processing does not buy robustness against catastrophic failure on small primaries.

## Background

Head and neck cancer (HNC) is the seventh most common malignancy worldwide, with more than 890 000 new cases and 450 000 deaths annually [1]. Definitive or postoperative radiotherapy, frequently combined with concurrent chemotherapy, is a mainstay of curative treatment. Accurate delineation of the gross tumour volume (GTV) — comprising the primary tumour (GTVp) and involved lymph nodes (GTVn) — directly determines target coverage, dose conformity to organs at risk, and ultimately local control and toxicity. Inter-observer variability of manual GTV delineation is well documented, with reported inter-expert DSC in the 0.5 – 0.8 range and volume differences of 30 – 60 % [2, 3].

Fused FDG-PET/CT has become the reference imaging modality for HNC target delineation, combining CT anatomy with ^18^F-FDG metabolic contrast, following ESTRO-ACROP consensus guidelines [4]. Deep-learning-based auto-segmentation on multi-modal PET/CT is therefore an active area of research. The HECKTOR (HEad and neCK TumOR segmentation and outcome prediction) MICCAI challenge, held annually since 2020, has become the international benchmark for automatic GTV segmentation in HNC [5, 6, 7]. The 2025 edition (Task 1) provides 680 training cases from eight international centres and requires the more demanding *simultaneous* segmentation of GTVp and GTVn.

The nnU-Net framework [8], which auto-configures preprocessing, architecture and training on a per-dataset basis, has become the *de facto* baseline for medical image segmentation. Several top-ranking HECKTOR submissions have been nnU-Net variants [9, 10]. Subsequent work has moved in two directions: higher-capacity, transformer-driven convolutional designs [17], and more rigorous re-validation of the framework itself [16]. However, nnU-Net auto-derived architectures for HN PET/CT typically contain 80 – 100 M parameters (verified 88 M in our configuration) and rely on 5-fold ensembling for optimal performance, which imposes non-trivial compute and inference-time constraints in clinical radiotherapy departments equipped with a single planning workstation. Custom, hand-designed lightweight U-Net variants remain of interest for those reasons — reduced computational footprint, faster single-model inference, and easier maintenance in a clinical IT environment [11, 12].

Comparisons between such custom pipelines and framework baselines are, however, almost always reported end-to-end: a tuned custom pipeline (network + thresholding + morphology + component filtering) is placed against a framework’s default output, and any difference is attributed to the architecture. If the post-processing stage carries a substantial fraction of the performance, that attribution is wrong, and the practical guidance derived from it — “build a smaller network” — is misdirected.

### Contribution

This study contributes:

1. **A three-arm paired design that isolates post-processing from architecture** on HECKTOR 2025 Task 1, using an identical training/validation split for both networks: MiniUNet3D raw, MiniUNet3D post-processed, and nnU-Net v2 raw.
2. **A quantification of the relative contribution of each factor**, showing that in this task the inference-time post-processing stage recovers 79 % of the gap between the two trained models, so that the difference surviving it is three to four times smaller than the post-processing contribution itself.
3. **Statistically-controlled inference**: paired Wilcoxon tests with bootstrap confidence intervals, Bonferroni and BH-FDR correction over the metric family, and effect sizes reported alongside every p-value.
4. **An explicit catastrophic-failure endpoint** analysed with an exact McNemar test, distinguishing an accuracy difference from a safety difference.
5. **Volume-stratified subgroup analysis using *native* ground-truth tumour volumes** (invariant to per-model resampling), together with failure-mode characterisation by tumour volume and originating centre.

## Methods

The study is reported in accordance with the CLAIM (Checklist for AI in Medical Imaging) recommendations [26]; the completed 42-item checklist, including the two items this study does not meet, is provided as Additional file 1. Random seed 42 was used for all splits and training initialisation to ensure reproducibility.

### Dataset

The HECKTOR 2025 MICCAI Challenge Task 1 training set was used [13]. It comprises 680 patients with histologically confirmed HNC from eight international centres (MDA Houston, CHUM, CHUP, CHUS, HGJ, HMR, USZ and additional contributors), each imaged with co-registered ^18^F-FDG PET/CT and provided with expert manual delineations of GTVp and GTVn as multi-class masks (background = 0, GTVp = 1, GTVn = 2), named per the AAPM TG-263 nomenclature [14].

Because the HECKTOR 2025 test set is retained by the challenge organisers for the official leader-board evaluation, we defined our own fixed patient-level 85 / 15 split of the 680 released training cases: **578 training** and **102 internal validation** cases, stratified by centre. The exact same split was used for both MiniUNet3D and nnU-Net v2 training. Validation cohort composition (n = 102): MDA (n = 58, 56.9 %), CHUS (n = 13), CHUP (n = 12), HGJ (n = 10), CHUM (n = 6), USZ + HMR (n = 3).

#### Cohort definition per target

Four validation cases have no primary tumour in the reference (N/A) and nine are node-negative (N0). A prediction of an empty mask against an empty reference scores DSC = 1.000, which rewards a model for correctly predicting nothing and inflates the arm that tends to predict empty masks. These cases were therefore excluded from the corresponding target throughout — **descriptive statistics, paired tests and failure analysis alike**. The effective cohorts are **n = 98 for all GTVp metrics** and **n = 93 for all GTVn metrics**; macro-Dice, which averages over both foreground classes, retains all 102 cases. This filtering also explains a small (∼0.005) upward shift in GTVp median DSC relative to a preliminary conference presentation of the same model, which used the full 102-case denominator.

### Preprocessing

All volumes were resampled to a target isotropic voxel spacing of 1.5×1.5×1.5 mm using B-spline interpolation for images and nearest-neighbour for labels. CT intensities were clipped to [–1024, 3071] HU and z-score normalised per case. PET intensities provided by HECKTOR are in decay-corrected activity units (Bq/mL); we converted them to body-weight standardised uptake values (SUV, following EANM 2015 guidelines [27]) using per-case injected dose, weight, and acquisition delay from the DICOM headers, and then applied per-case z-score normalisation.

### MiniUNet3D architecture

MiniUNet3D is a symmetric encoder-decoder 3D U-Net [19, 20] with feature maps of [32, 64, 128, 256, 384] channels across four encoder stages and a bottleneck. Each convolutional block consists of two sequential 3×3×3 convolutions with instance normalisation and LeakyReLU (0.1). Down-sampling is 3D max-pooling (2 x 2 x 2); up-sampling is 3D transposed convolution (2×2×2). Skip connections concatenate encoder and decoder feature maps at each resolution. The output layer produces three logits with soft-max activation. Total parameters: **18 332 547 (18.3 M)**, counted from the released checkpoint.

### Training

The model was trained end-to-end on 144^3^-voxel patches with a batch size of 4 on a single NVIDIA RTX 6000 Ada GPU (48 GB VRAM). The loss was a weighted sum of Dice loss and voxel-level cross-entropy (equal weights). Optimisation used AdamW [21] with mixed-precision training (AMP) throughout [22]. **A provenance caveat applies to the optimiser configuration**: the training run predates the archival of this project’s code, and the three surviving records disagree on the weight-decay value and the learning-rate schedule (the archived notebook released with this work specifies weight decay 10^−4^ with plateau-based decay; the optimiser state stored in an intermediate checkpoint records 10^−2^). We therefore report only what can be verified from the released artefacts and flag the remainder as not reconstructible, rather than stating a configuration we cannot evidence.

On-the-fly augmentations were applied with per-sample independent probability: random rotations ±10° (p = 0.5), random flips along each spatial axis (p = 0.5 per axis) and additive Gaussian noise (mean 0, SD 0.01; p = 0.15).

The best model was selected by monitored macro-Dice on a held-out development subset (n = 52) drawn from the 578-case training pool, and is the checkpoint released with this work: **epoch 49, development Dice 0.722**, both read directly from that checkpoint. The surviving per-epoch checkpoint series extends to epoch 75, so the run length exceeded the selected epoch by a comfortable margin; we do not state a nominal epoch budget, because the records that survive do not agree on one. That subset was used for model selection and for post-processing hyperparameter search, and never for weight updates, so **MiniUNet3D was fitted on 526 cases and used 52 for selection**, whereas nnU-Net v2 consumed the full 578 under its own internal scheme. This asymmetry in training budget is revisited in the Limitations.

### Inference and adaptive post-processing

Sliding-window inference with 50 % overlap and Gaussian re-weighting produced dense probability maps for each output class. Test-time augmentation (TTA) applied seven flip combinations along the sagittal, coronal and axial axes and averaged the resulting probability maps. **nnU-Net v2 applies mirroring TTA by default and was run in that default configuration, so both arms include test-time augmentation**; the arms are symmetric in this respect.

Raw probability maps were processed by an adaptive post-processing pipeline whose hyperparameters were selected on the same **held-out development subset (n = 52) drawn from the 578-case training pool and never used for training weights**, and then locked before evaluation on the internal validation cohort. The final pipeline (v9, after nine internal iterations) applies (i) per-class thresholding at 0.50 with adaptive per-case downward relaxation for low-confidence cases, (ii) morphological opening with a 3×3×3 structuring element to remove speckle and (iii) 3D connected-component analysis retaining components larger than 0.10 mL (GTVp) and 0.05 mL (GTVn). The 0.10 / 0.05 mL thresholds were selected on the development subset to minimise false-positive components without discarding real small primaries. Mean end-to-end inference time on the same GPU was **34 s per case**, including TTA and post-processing.

### nnU-Net v2 baseline

For direct methodological comparison we trained **nnU-Net v2** [8] with the auto-configured `3d_fullres` plan and the `nnUNetTrainer_500epochs` trainer variant, on the identical 578-case training set, and predicted the same validation cohort. Default hyperparameters were used; no further tuning was applied. The auto-derived architecture contains **88 215 055 parameters (88.2 M)**, counted from the released checkpoint — approximately 4.8x the MiniUNet3D count, so MiniUNet3D uses ∼21 % of the nnU-Net parameter budget. A single fold trained on the paper’s fixed 578 / 102 split was used, to preserve a like-for-like *single-model* comparison. Mean end-to-end inference time was 78 s per case on the same GPU (about 2.3× slower than MiniUNet3D).

nnU-Net v2 predictions were evaluated as produced by the framework’s default inference. To establish whether this places the baseline at a disadvantage, we additionally ran the framework’s own post-processing selection step, `nnUNetv2_determine_postprocessing`, on the fold’s validation output. **The procedure selected an empty post-processing configuration**: neither retaining only the largest foreground region nor retaining only the largest connected component per class improved the result, and forcing the latter *reduced* mean Dice for both targets (GTVp 0.704 to 0.698; GTVn 0.673 to 0.651; values from nnU-Net’s internal evaluation in its own preprocessed space and therefore not comparable to Table 2). Notably, this selection was performed on the evaluation cohort itself — nnU-Net’s standard procedure — which if anything favours the baseline, and it still yielded no operations. The framework’s native post-processing therefore has nothing to contribute on this dataset, and the arm C results stand as the framework’s best available configuration.

The comparison remains asymmetric in one respect, and the study is designed with three arms because of it: MiniUNet3D carries a bespoke adaptive pipeline developed over nine internal iterations, whereas nnU-Net offers only the built-in connected-component step just shown to be inert here. The raw-versus-raw contrast (arm A vs arm C) removes post-processing as a confounder and is therefore the cleaner of the two; we nevertheless refer to it throughout as a comparison of the two *trained models*, not of their architectures, because the arms also differ in training budget (a run whose surviving checkpoints reach epoch 75, on 526 cases, versus 500 epochs on 578). Attributing the resulting gap to architecture alone would not be warranted. The post-processed-versus-raw contrast (arm B vs arm C) reflects deployed pipelines. Both are reported.

Two further asymmetries are declared: the training budgets differ substantially — the MiniUNet3D run’s surviving checkpoints reach epoch 75 on 526 cases, against 500 epochs on 578 for nnU-Net v2 — and the recommended nnU-Net v2 deployment configuration is a 5-fold ensemble, which typically outperforms any single fold [8, 16].

### Evaluation metrics

For each patient and each of the two target volumes we computed:

- **Dice similarity coefficient (DSC)**: 2 |*A* ∩ *B*| / (|*A*| + |*B*|), reported as median, IQR, and mean ± SD.
- **Surface DSC at 2 mm tolerance (sDSC@2mm)**: symmetric mean of the fractions of the predicted and reference surfaces within a 2 mm tolerance of the other surface [15].
- **95th-percentile Hausdorff distance (HD95)** in mm. HD95 is undefined when either mask is empty; such pairs are dropped pairwise, which reduces N for HD95 rows and does so more severely in the raw arms.

Metrics were selected in line with the Metrics Reloaded framework for medical image analysis [25]. **Ground-truth tumour volume was computed once per case on the native reference file** (before any per-model resampling) and used consistently across both models for the volume-stratified analysis, ensuring that per-bin subgroups contain the same patients regardless of the model being evaluated. Volume bins were: <2, 2 – 5, 5 – 10, 10 – 20, 20 – 50 and > 50 mL. Segmentation *failures* were defined as DSC <0.01 (essentially empty overlap), with a sensitivity analysis at DSC <0.20 reported inline in the failure-mode section.

### Statistical analysis

Paired per-patient comparison used the two-sided **Wilcoxon signed-rank test** on the per-patient difference. **Bootstrap 95 % confidence intervals for the mean paired difference** were computed with 10 000 resamples (seed 42). **Multiple-testing correction** was applied within each comparison over its family of metrics using both **Bonferroni** and the **Benjamini–Hochberg false discovery rate** (BH-FDR) [28]. Effect sizes were reported as **Cohen’s d** <u>fo</u>r the paired difference (*d_z_* = mean difference / SD of the difference) and **Wilcoxon** 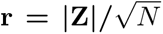. Catastrophic failure rates were compared with the **exact (binomial) McNemar test** on discordant pairs [29]. Pearson correlation was used for continuous predictors (GTVp volume vs DSC).

A two-sided adjusted p-value <0.05 was considered statistically significant. Where a comparison fails to reject the null, we do **not** interpret this as evidence of equivalence: no non-inferiority margin was pre-specified, and with n = 98 paired cases at 80 % power the minimum detectable paired effect is approximately *d_z_* = 0.29, so effects smaller than that would not be reliably detected [23, 24]. All analyses were performed in Python 3.11 with SimpleITK 2.5, SciPy 1.11, statsmodels 0.14, NumPy 1.26 and pandas 2.3.

## Results

### Trained models with post-processing matched off (arm A vs arm C)

With post-processing absent from both arms — MiniUNet3D thresholded at a fixed 0.5 with no morphology and no component filtering, nnU-Net v2 at its default output — the two trained models are far apart (Table 1). We label this a model-level rather than an architectural comparison because the arms differ in training budget as well as in architecture.

**Table 1.** Comparison of the two trained models with post-processing matched off: MiniUNet3D raw vs nnU-Net v2 raw. Values are median (mean). Delta mean is the paired per-patient mean difference (MiniUNet3D minus nnU-Net v2); 95 % CI from a 10 000-sample bootstrap; p_raw is two-sided Wilcoxon signed-rank; p_Bonf and p_FDR are corrected over this family of four tests.

| Metric | N | MiniUNet3D<br>raw | nnU-Net<br>v2 | Delta mean [95 %<br>CI] | p_raw | p_Bonf | p_FDR | d | r |
| --- | --- | --- | --- | --- | --- | --- | --- | --- | --- |
| Dice — | 98 | 0.592 | **0.799** | −0.244 [−0.300, | <0.001 | <0.001 | <0.001 | −0.88 | 0.73 |
| GTVp |  | (0.480) | (0.724) | −0.191] |  |  |  |  |  |
| Dice — | 93 | 0.598 | **0.774** | −0.246 [−0.306, | <0.001 | <0.001 | <0.001 | −0.82 | 0.69 |
| GTVn |  | (0.477) | (0.723) | −0.186] |  |  |  |  |  |
| HD95 — | 74 | 5.41 (8.05) | **3.18** | −4.22 [−18.93, | <0.001 | <0.001 | <0.001 | −0.07 | 0.50 |
| GTVp (mm) |  |  | (12.26) | +3.84] |  |  |  |  |  |
| HD95 — | 80 | 15.98 | 11.42 | −18.33 [−40.84, | 0.522 | 1.000 | 0.522 | −0.20 | 0.07 |
| GTVn (mm) |  | (27.02) | (45.34) | −0.50] |  |  |  |  |  |

nnU-Net v2 is superior on both overlap metrics with **large effect sizes** (Cohen’s d = –0.88 and –0.82) and adjusted p-values below 0.001. The reduced N in the HD95 rows (74 and 80 of 98/93) is itself informative: raw MiniUNet3D produces empty predictions often enough that the distance metric becomes undefined in roughly a quarter of cases.

### Deployed pipelines (arm B vs arm C)

Table 2 repeats the comparison with the adaptive post-processing pipeline active on MiniUNet3D, which is the configuration a department would deploy.

**Table 2.** Post-processed MiniUNet3D vs nnU-Net v2. Conventions as in Table 1; correction is over this family of seven tests.

| Metric | N | MiniUNet3D<br>v9 | nnU-Net<br>v2 | Delta mean [95 %<br>CI] | p_raw | p_Bonf | p_FDR | d | r |
| --- | --- | --- | --- | --- | --- | --- | --- | --- | --- |
| macro-Dice | 102 | 0.702 | 0.725 | −0.028 [−0.062, | 0.010 | 0.067 | **0.022** | −0.16 | 0.26 |
|  |  | (0.654) | (0.682) | +0.006] |  |  |  |  |  |
| Dice — GTVp | 98 | **0.800** | 0.799 | −0.051 [−0.092, | 0.113 | 0.790 | 0.174 | −0.26 | 0.16 |
|  |  | (0.672) | (0.724) | −0.014] |  |  |  |  |  |
| Dice — GTVn | 93 | 0.738 | **0.774** | −0.062 [−0.102, | 0.006 | **0.041** | **0.022** | −0.32 | 0.29 |
|  |  | (0.661) | (0.723) | −0.023] |  |  |  |  |  |
| sDSC @ 2 mm | 98 | 0.859 | 0.883 | −0.040 [−0.078, | 0.124 | 0.868 | 0.174 | −0.21 | 0.15 |
| — GTVp |  | (0.768) | (0.809) | −0.004] |  |  |  |  |  |
| sDSC @ 2 mm | 93 | 0.771 | **0.822** | −0.043 [−0.078, | 0.007 | **0.049** | **0.022** | −0.25 | 0.28 |
| — GTVn |  | (0.751) | (0.794) | −0.008] |  |  |  |  |  |
| HD95 — | 90 | 3.96 (5.69) | 3.51 | −7.42 [−19.78, | 0.574 | 1.000 | 0.670 | −0.14 | 0.06 |
| GTVp (mm) |  |  | (13.11) | −0.33] |  |  |  |  |  |
| HD95 — | 93 | 12.15 | 11.81 | −15.99 [−37.11, | 0.778 | 1.000 | 0.778 | −0.16 | 0.03 |
| GTVn (mm) |  | (27.30) | (43.30) | +3.92] |  |  |  |  |  |

On the primary radiotherapy target the two pipelines are statistically indistinguishable (GTVp DSC 0.800 vs 0.799; p_raw = 0.113, adjusted p = 0.790). On nodal disease nnU-Net v2 retains an advantage that **survives Bonferroni correction** (DSC adjusted p = 0.041; sDSC@2mm adjusted p = 0.049), with small-to-moderate effect sizes (d = –0.32 and –0.25). HD95 differences are not significant in either direction; the bootstrap CI for GTVp HD95 excludes zero, but this reflects the mean of a heavily right-skewed distribution driven by a few nnU-Net outlier cases and should not be read as a robust advantage.

**Figure 1:**
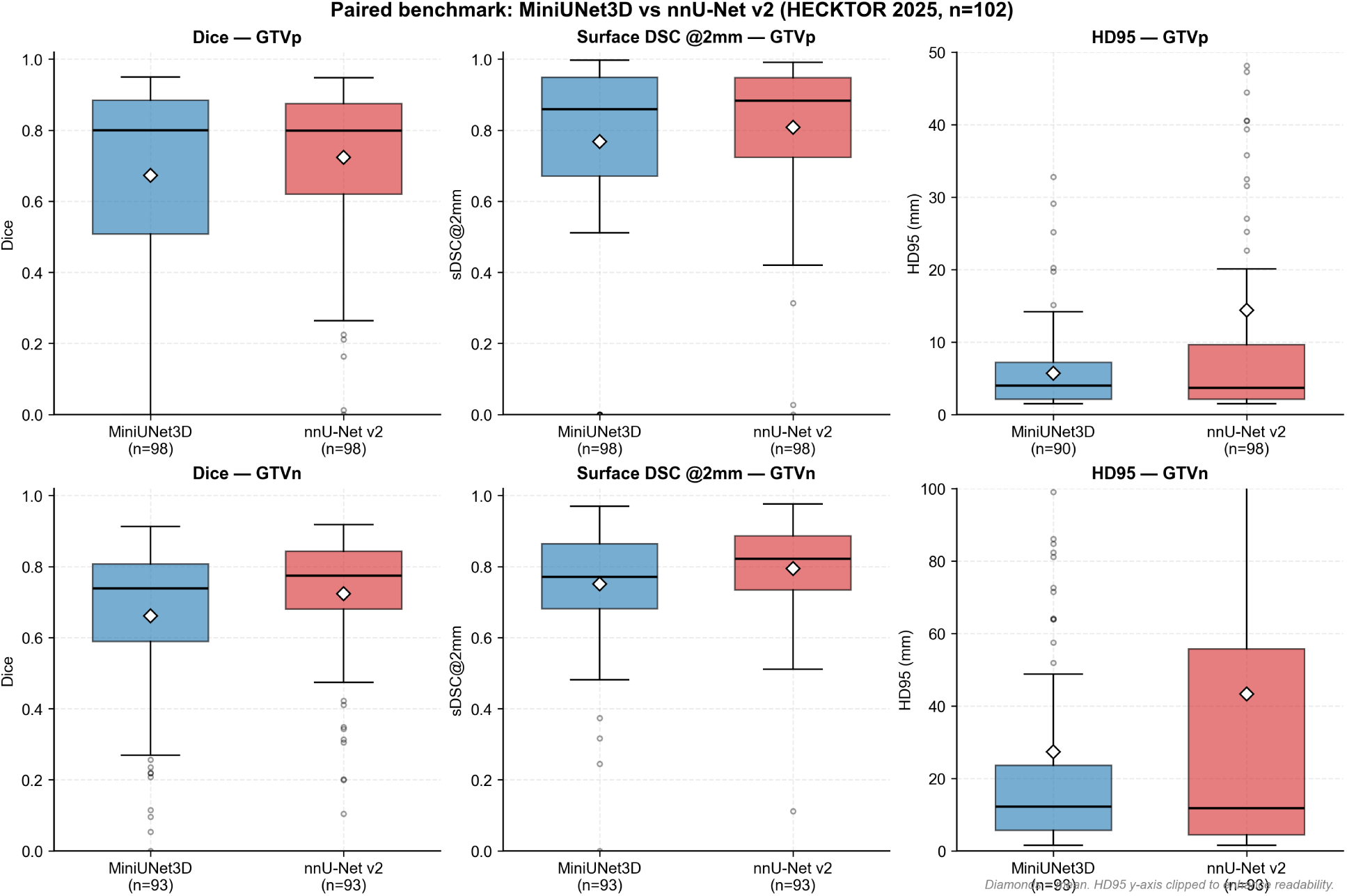
Paired per-patient distributions of DSC, sDSC@2mm and HD95 for GTVp (top row) and GTVn (bottom row), post-processed MiniUNet3D vs nnU-Net v2. Diamonds mark the mean; centre lines the median; whiskers 1.5 x IQR. HD95 y-axes clipped for readability.

For clinical context, published inter-expert DSC for manual HN GTV delineation lies in the range 0.5 – 0.8 [2, 3]; both post-processed pipelines sit at or above the upper end of that reference range for GTVp, consistent with auto-segmentation approaching the ceiling imposed by human labelling variability.

### Post-processing ablation (arm A vs arm B)

Table 3 isolates the post-processing contribution within MiniUNet3D.

**Table 3.**
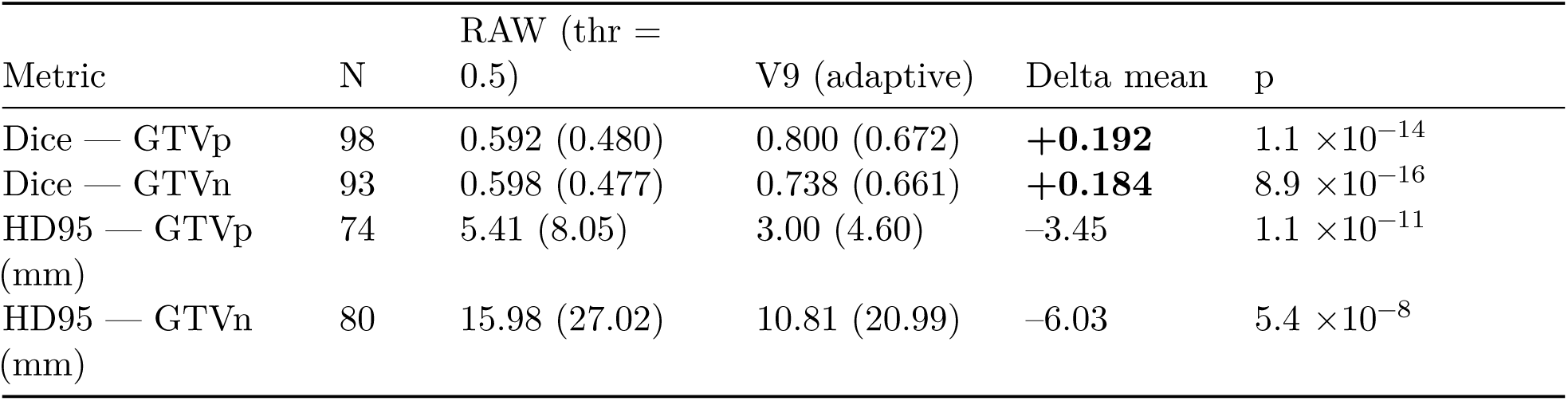
Post-processing ablation on MiniUNet3D: raw (fixed threshold 0.5) vs adaptive v9 pipeline, paired on the same cases. Values are median (mean); Delta mean is V9 minus RAW; p is two-sided Wilcoxon signed-rank.

The comparison of magnitudes is the central result of this study, and it must be stated with care. The model-level difference with post-processing matched off is **0.244 DSC** (Table 1). Post-processing moves MiniUNet3D by **+0.192 DSC**, that is **79 %** of it: the post-processing stage is therefore *slightly smaller* than the model gap, not larger. What changes is the residual: after post-processing, the difference between the two pipelines is **–0.051** on GTVp and at most **–0.062** on any metric in Table 2, so the post-processing contribution is **three to four times** whatever model-level difference survives it. The practical statement is that post-processing does not outweigh the choice of model — it very nearly cancels it.

Because the pipeline hyperparameters were selected on a training-pool development subset (n = 52) and locked before evaluation, this gain reflects generalisation of the post-processing rather than tuning on the reported evaluation cohort.

### Volume-stratified analysis (GTVp)

Ground-truth tumour volumes were computed from the native reference files, ensuring identical per-bin patient composition across methods. GTVp DSC was moderately correlated with tumour volume in both models (Pearson r = 0.39 for MiniUNet3D and 0.41 for nnU-Net v2; both p <0.001).

**Table 4.** GTVp performance stratified by native ground-truth tumour volume, post-processed MiniUNet3D vs nnU-Net v2 (paired subset, n = 98).

| GT volume | $n$ | MiniUNet3D v9 median | nnU-Net v2 median | Delta median |
| --- | --- | --- | --- | --- |
| <2 mL | 7 | 0.391 | **0.449** | -0.058 |
| 2 – 5 mL | 21 | 0.640 | 0.664 | -0.023 |
| 5 – 10 mL | 27 | 0.654 | **0.765** | -0.111 |
| 10 – 20 mL | 17 | 0.830 | 0.872 | -0.042 |

| GT volume | n | MiniUNet3D v9 median | nnU-Net v2 median | Delta median |
| --- | --- | --- | --- | --- |
| 20 – 50 mL | 23 | 0.886 | 0.886 | +0.000 |
| >50 mL | 3 | 0.884 | 0.838 | +0.046 |

nnU-Net v2 was numerically better or equivalent in all volume bins except the very-large (>50 mL, n = 3) bin, with its largest advantage in the 5 – 10 mL range. No individual bin comparison reached significance after correction (small per-bin n). That post-processed MiniUNet3D matches nnU-Net v2 on the overall GTVp median while trailing in most bins reflects a redistribution of case-level errors rather than per-bin parity.

### Failure modes and catastrophic failure rate

**Table 5.** Catastrophic GTVp failures (DSC <0.01) among the 98 valid-primary cases, with exact McNemar tests against nnU-Net v2 on discordant pairs.

| Arm | Failures / 98 | Rate | Discordant vs nnU-Net (arm-only :<br>nnU-Net-only) | McNemar<br>exact p |
| --- | --- | --- | --- | --- |
| MiniUNet3D<br>raw | 25 | 25.5 % | 24 : 0 | $< 0.001$ |
| MiniUNet3D<br>v9 | 8 | 8.2 % | 7 : 0 | **0.016** |
| nnU-Net v2 | 1 | 1.0 % | — | — |

At the alternative sensitivity threshold DSC <0.20 the counts are 33, 10 and 3 respectively, preserving the ordering.

Post-processing removes two thirds of the raw failures (25→8) but does not close the gap to nnU-Net v2: the eight-fold difference in catastrophic failure rate remains statistically significant (McNemar exact p = 0.016, seven discordant pairs all in the same direction). All MiniUNet3D failures occurred in cases with ground-truth tumour volume ≤ 7.63 mL. They were distributed across **5 of the 7 validation centres** (MDA n = 4, CHUM n = 1, CHUS n = 1, HGJ n = 1, USZ n = 1), ruling out a single-centre scanner or protocol artefact; the single nnU-Net v2 failure (case USZ-008, 3.14 mL) also failed under MiniUNet3D. GTVn failures were rare in both models (<2 %).

Figure 4 shows representative axial slices for a best case, a typical case and a small-primary failure case where post-processed MiniUNet3D returned an empty prediction while nnU-Net v2 achieved DSC = 0.75.

**Figure 2:**
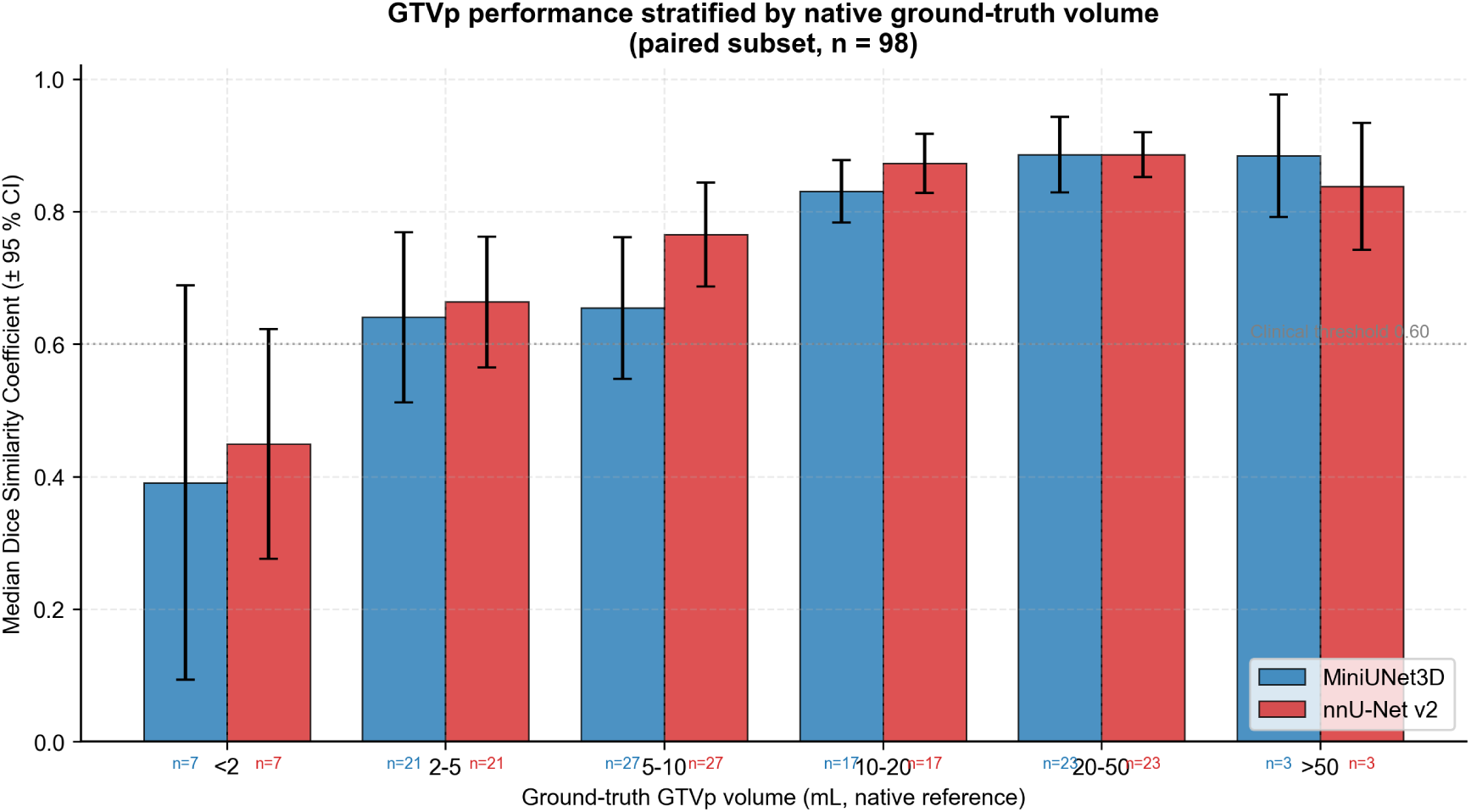
GTVp median DSC stratified by native ground-truth tumour volume, with 95 % confidence intervals derived from the standard error of the mean per bin (approximate; small per-bin samples). Both methods are evaluated on identical patient subsets per bin.

**Figure 3:**
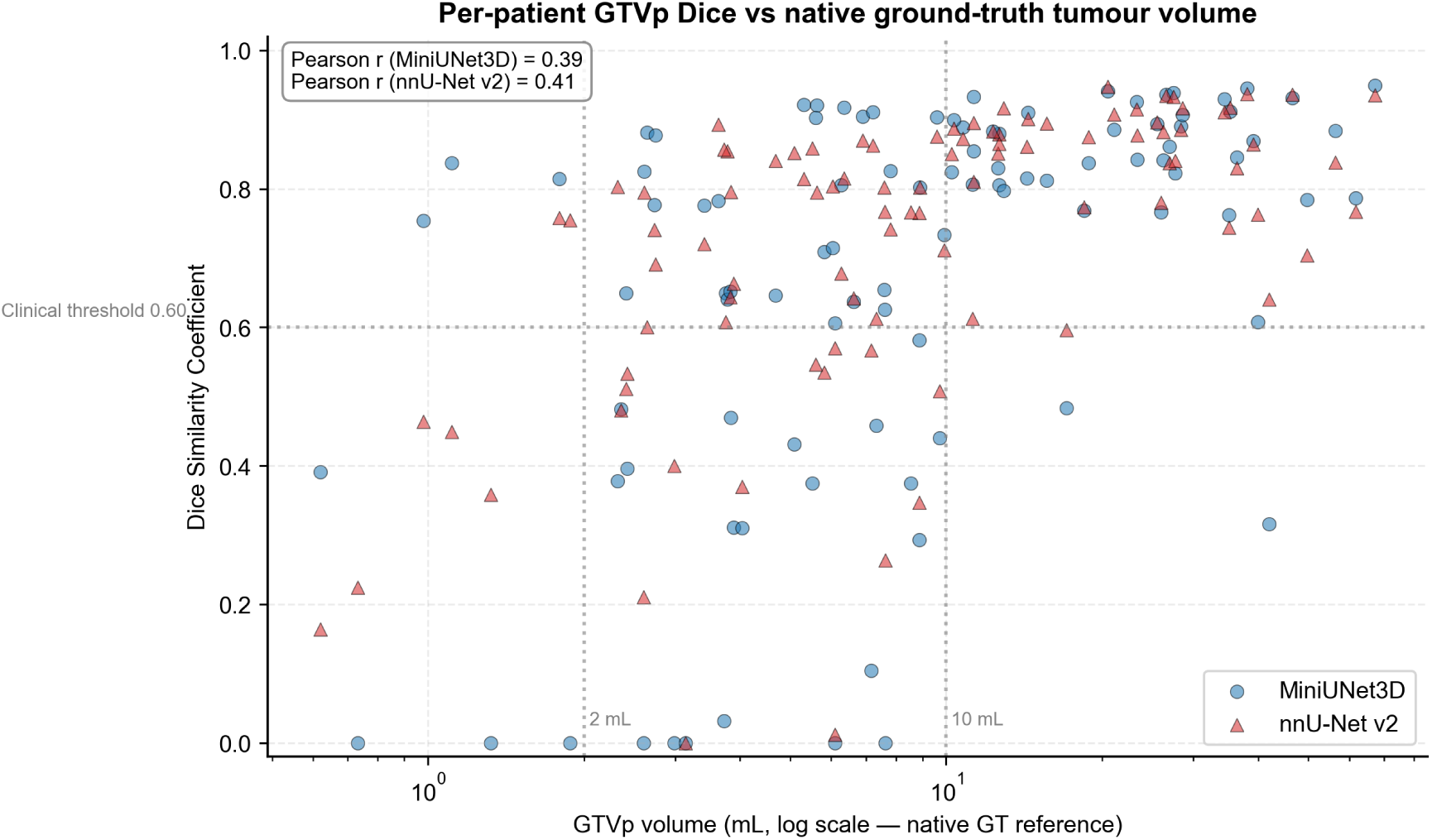
Per-patient GTVp DSC vs native tumour volume (log scale). Vertical dotted lines at 2 mL and 10 mL delimit the low- and mid-volume regimes; horizontal line at 0.60 marks the clinical acceptability threshold typically used in HN GTV auto-segmentation studies.

**Figure 4:**
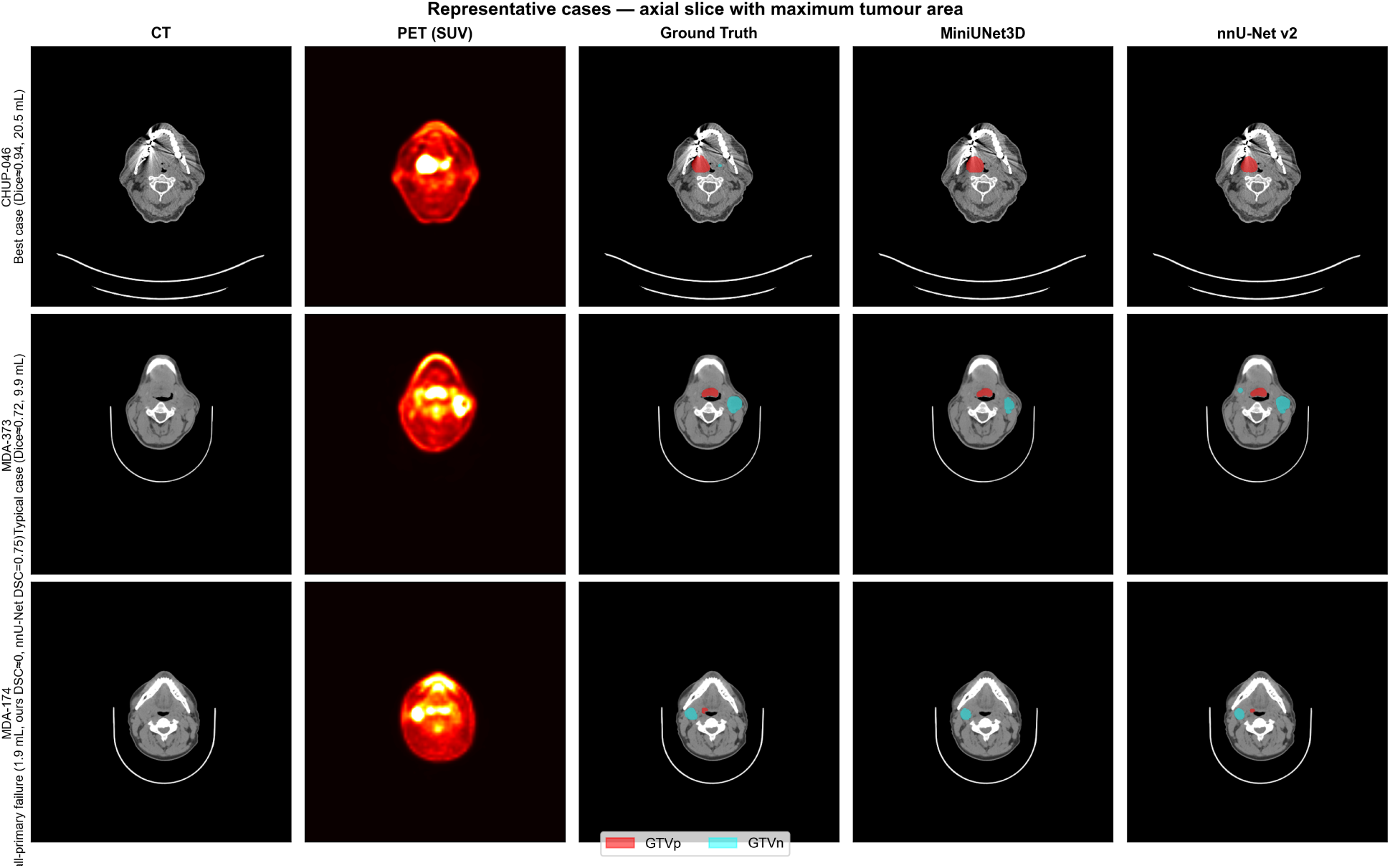
Representative cases (axial slice with maximum tumour area). Rows top-to-bottom: best case (CHUP-046, 20.5 mL, both methods excellent), typical case (MDA-373, 9.9 mL, both methods clinically acceptable), and small-primary failure case (MDA-174, 1.9 mL, MiniUNet3D empty vs nnU-Net v2 DSC = 0.75). Columns: CT, PET (SUV heatmap), Ground Truth overlay on CT, MiniUNet3D prediction overlay on CT, nnU-Net v2 prediction overlay on CT. Red = GTVp; cyan = GTVn.

### Qualitative examples

Figure 4 shows three cases chosen by hand, which is a selection a reader is entitled to distrust. Figures 5 and 6 therefore show a gallery drawn by a rule fixed in advance: the 98 GTVp-evaluable cases were ordered by the post-processed DSC of arm B and 12 of them were taken at evenly spaced positions along that ordering, so the panel spans the whole performance range (DSC 0.000 to 0.949) and cannot be curated. Each row shows the same axial slice under all three arms, with the per-case DSC printed beneath each prediction.

**Figure 5:**
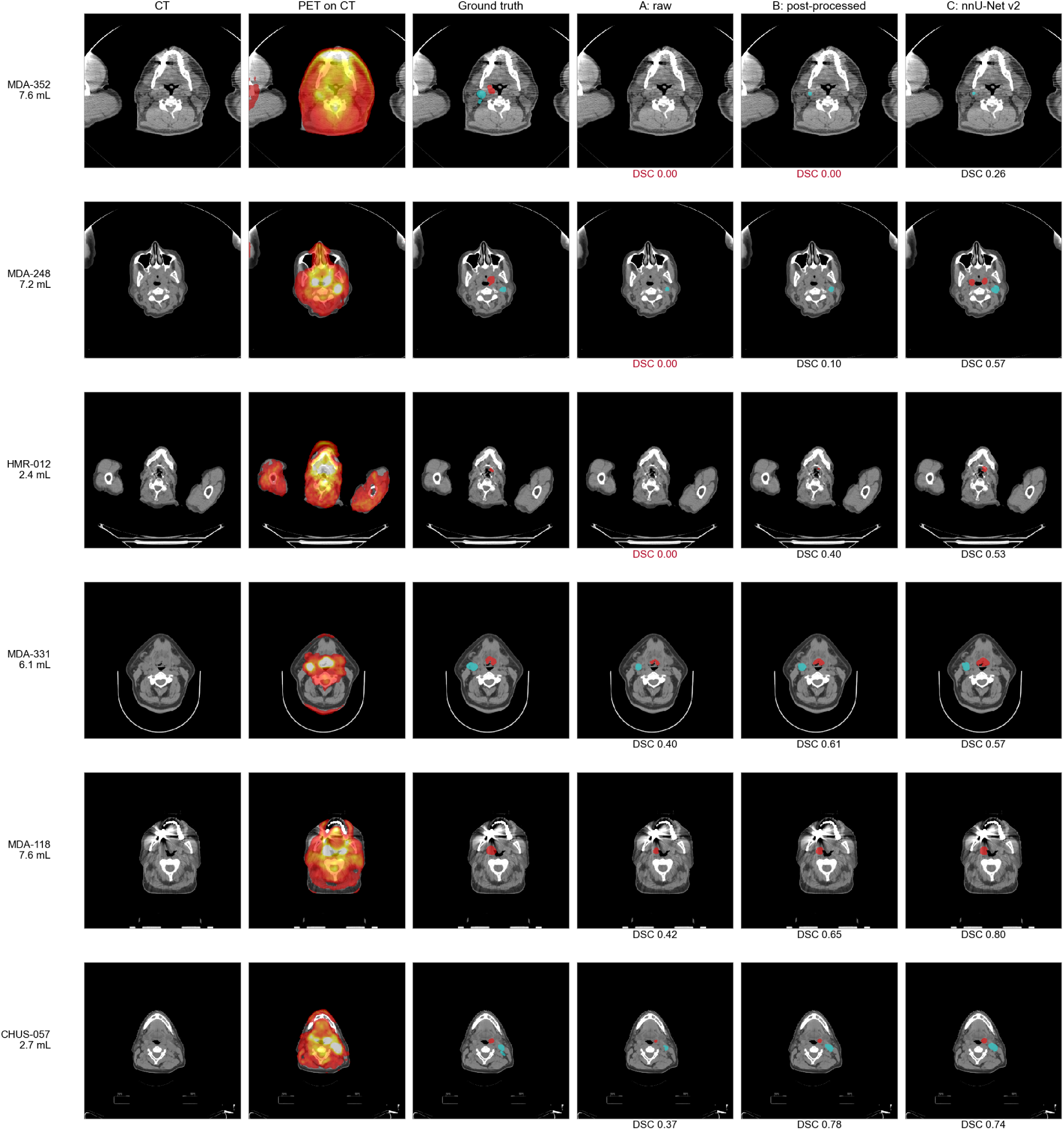
Qualitative gallery, part 1 of 2 — the 6 lower-performing of the 12 cases selected. Columns: CT; PET fused on CT (masked below 25 % of the per-slice maximum, so that the point-spread blur of PET does not obscure the anatomy it is registered to); ground truth; arm A (raw, fixed 0.5 threshold); arm B (adaptive post-processing); arm C (nnU-Net v2). Red = GTVp, cyan = GTVn. Row labels give the case identifier and its native ground-truth GTVp volume; DSC beneath each prediction is the per-case value from the same file used for all statistics, with empty contours in red. Cases were selected by the pre-stated rule described in the text, not chosen.

**Figure 6:**
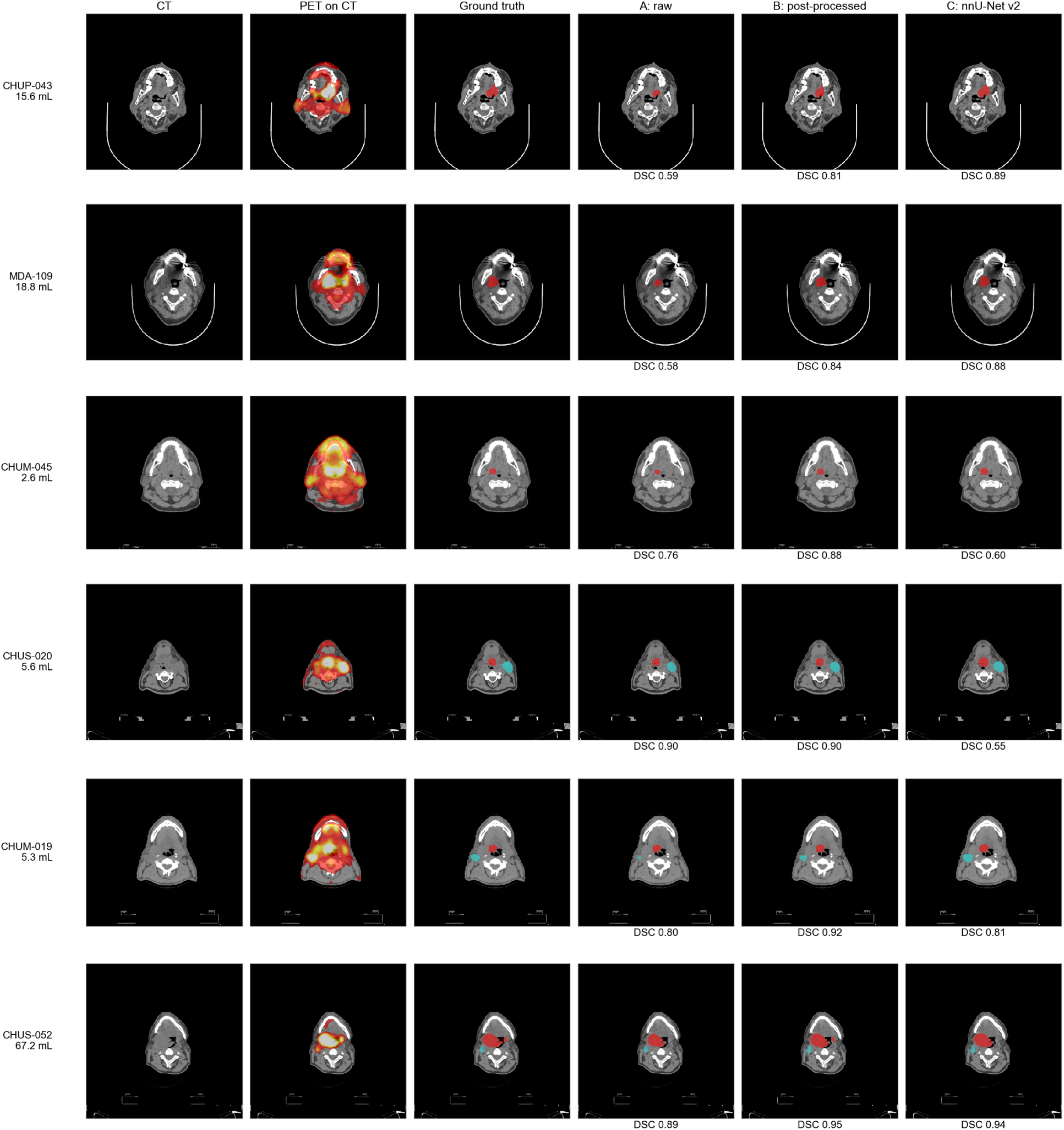
Qualitative gallery, part 2 of 2 — the 6 higher-performing cases. Columns, colours and labels as in the preceding figure.

The ablation is visible case by case: post-processing improves the primary contour in 10 of the 12 cases shown, and in 6 of them the post-processed pipeline also exceeds nnU-Net v2 on that case — a proportion that matches the cohort as a whole, where arm B exceeds arm C in 47 of the 98 evaluable cases, as expected of two pipelines that are statistically indistinguishable on this endpoint. The one row where arm B still returns an empty contour (MDA-352, 7.6 mL) and the one where it recovers only a fragment (MDA-248, 7.2 mL, DSC 0.10) both fall inside the ≤ 7.63 mL regime in which every catastrophic failure in this cohort occurred.

## Discussion

We benchmarked a lightweight custom 3D U-Net against the auto-configured nnU-Net v2 on HECKTOR 2025 Task 1 using a three-arm paired design that separates the contribution of the network from that of the inference-time post-processing. Four findings shape the interpretation.

*First*, **the two trained models are not close.** With post-processing matched off, nnU-Net v2 outperforms MiniUNet3D by 0.244 median DSC on GTVp and 0.246 on GTVn, with large effect sizes (d = –0.88 and –0.82) and adjusted p-values below 0.001. How much of that gap is architecture and how much is the 4-fold difference in training budget cannot be separated by this study, and we do not attempt to. What the comparison does establish is that any account beginning from the deployed pipelines and concluding that “a 18 M-parameter network matches an 88 M-parameter framework” is describing the post-processing, not the models.

*Second*, **post-processing recovers 79 % of that gap.** A locked adaptive pipeline — confidence-relaxed thresholding, morphological opening and small-component filtering — adds 0.192 DSC to MiniUNet3D on GTVp. We are deliberate about what this does and does not say: 0.192 is *less* than the 0.244 model-level difference, so post-processing does not outweigh the choice of model. It nearly cancels it, and what survives (0.051 – 0.062) is three to four times smaller than the post-processing contribution. This has a direct practical reading for departments building their own tooling: a team that has already committed to a lightweight custom network — for deployment, maintenance or latency reasons — can recover most of what that choice costs at the inference stage, without retraining. It does not say that post-processing is a substitute for adopting the stronger model in the first place; that remains the single largest lever available. It also has a methodological reading for the field: **end-to-end pipeline comparisons without a post-processing ablation are not architecture comparisons**, and the ablation is cheap relative to the training it contextualises.

The gain is specific to the adaptive pipeline rather than to post-processing in general. nnU-Net’s own connected-component selection, run on the evaluation cohort itself, returned an empty configuration and degraded both targets when forced (Methods) — so the recovery reported here comes from confidence-relaxed thresholding and volume-aware component filtering, not from generic clean-up that any framework supplies by default.

*Third*, **post-processing does not confer robustness.** It removes two thirds of the catastrophic failures (25→8 of 98) but leaves an eight-fold gap against nnU-Net v2 that survives an exact McNemar test (p = 0.016, seven discordant pairs, all in the same direction). This is the difference that matters most clinically: an empty contour on a planning study is a safety event requiring detection by the reviewing clinician, not a marginal loss of overlap. All failures concentrated in primaries ≤7.63 mL and spread across five of seven centres, so the mode is a property of the model rather than of an acquisition protocol, and would need to be addressed by architectural or data-augmentation strategies targeting low-volume, high-SUV lesion detection. Prospective clinical evaluation of auto-segmentation for oropharyngeal GTV on FDG-PET/CT has begun to be reported [18], and rare catastrophic failures of this kind are precisely what such evaluations must be powered to capture. The residual nodal-disease advantage of nnU-Net v2 (GTVn DSC and sDSC@2mm, both Bonferroni-significant) points the same way: the deeper auto-derived configuration handles the greater anatomical dispersion and shape variability of nodal disease better, and post-processing does not compensate for it.

*Fourth*, **the deployment trade-off is real but narrower than a pipeline-level comparison suggests.** Post-processed MiniUNet3D matches nnU-Net v2 on the primary endpoint (GTVp DSC 0.800 vs 0.799) at ∼21 % of the parameters and 34 s versus 78 s per case. For a department with a single planning workstation this remains a meaningful operational difference. But it is purchased with a higher catastrophic failure rate on small primaries and a small deficit on nodal disease, and it depends on a post-processing stage whose equivalent was never applied to the baseline.

### Relation to challenge performance

Our figures are not comparable to the HECKTOR leader-board and should not be read against it. The 2025 test set is retained by the organisers, so our evaluation uses an author-defined 15 % split of the released training data; the challenge ranks on an aggregated Dice computed over its own held-out cases, and its 2025 testing phase was still open when this manuscript was prepared (checked August 2026), so no final ranking exists to compare with. As an order-of-magnitude anchor only, the first edition of the challenge reported a best average DSC of 0.7591, against 0.6610 for the organisers’ baseline and 0.61 for inter-observer agreement [6] — on a task easier than the one addressed here, since it required the primary tumour alone on a single held-out centre rather than simultaneous GTVp and GTVn segmentation across eight. We report this to situate the reader, not as a benchmark: no shared cohort and no shared metric underlie the two sets of numbers, and treating them as commensurable would repeat precisely the attribution error this study was designed to expose.

We deliberately do not claim equivalence anywhere in this work. No non-inferiority margin was prespecified, and the study has approximately 80 % power to detect *d_z_* = 0.29; several of the observed effects lie below that threshold. A failure to reject is reported as such [23, 24].

### Limitations

- **The baseline received no bespoke post-processing.** nnU-Net’s own selection step was run and returned an empty configuration (Methods), so the framework’s native option is not the source of the difference; but no custom adaptive pipeline comparable to ours was developed for it, and we cannot exclude that one could be. The deployed-pipeline comparison (arm B vs arm C) should therefore be read as “a tuned custom pipeline against the framework’s best available configuration” — the situation a department actually faces — rather than as a controlled architectural contrast. The arm A vs arm C contrast is symmetric in post-processing and unaffected by this particular issue, though it carries its own confounder (below). Developing an equally-tuned pipeline for nnU-Net and repeating the comparison is the natural completion of this study.
- **Single-fold nnU-Net baseline.** The standard nnU-Net v2 deployment configuration is a 5-fold ensemble which typically outperforms any single fold [8, 16]. We used a single fold to preserve a single-model comparison. A 5-fold ensemble would be expected to widen, not narrow, the gaps reported here.
- **Asymmetric training budgets, and what that forbids us from claiming.** MiniUNet3D used 526 cases (52 reserved for model selection and post-processing tuning) and its selected checkpoint is epoch 49, with the surviving checkpoint series reaching epoch 75; nnU-Net v2 ran 500 epochs on 578 cases. Compute budget was not controlled, and the direction of the asymmetry favours nnU-Net v2. **This is why we describe arm A vs arm C as a comparison of two trained models rather than of two architectures throughout**: the 0.244 gap confounds architecture with training budget, and this study cannot apportion it. Retraining MiniUNet3D at matched budget is the experiment that would separate the two, and it may well narrow the gap; the post-processing finding, which is measured within a single model (arm A vs arm B), is unaffected either way.
- **No external validation, and public alternatives are less independent than they appear.** Validation was performed on our internally-held 102-case validation subset of the HECKTOR 2025 training set. We note for readers planning similar work that the largest public FDG-PET/CT head and neck collection carrying both primary and nodal GTV contours, TCIA Head-Neck-PET-CT, comprises 298 patients from four Québec institutions (CHUM, CHUS, HGJ, HMR) — the same four centres that contribute 201 of the 680 HECKTOR 2025 Task 1 cases. Using it as an external set without exact patient-level de-overlap would be circular rather than independent. Genuinely independent external validation therefore requires a cohort outside this lineage; it is planned in collaboration with the Radiation Oncology Department of Hospital Universitario Virgen del Rocío, Sevilla, Spain, and will be reported separately.
- **Centre imbalance.** MDA contributes 58 of 102 validation cases (56.9 %) while USZ and HMR together contribute 3. Aggregate results are therefore weighted towards a single centre, and no leave-one-centre-out sensitivity analysis was performed.
- **Small subgroup sizes.** Volume-stratified subgroups contain 3 – 27 patients each and no per-bin comparison reaches significance after correction. The failure-mode analysis rests on small discordant counts (7 pairs), and the McNemar result should be confirmed on an independent cohort.
- **No stratification by primary tumour site.** HN cancer comprises anatomically and biologically distinct sites (oropharynx, hypopharynx, larynx, oral cavity, nasopharynx) with different SUV patterns and expected segmentation difficulty. Per-site clinical metadata for the HECKTOR 2025 training set was not readily extractable in a form matching our validation subset; a per-site subgroup analysis is planned in the external-validation follow-up study.
- **No dosimetric impact evaluation.** The clinical criterion of interest for radiotherapy planning is the equivalence of the resulting dose distributions, not solely the DSC of the target contour. A dosimetric pilot (dose recalculation from auto-contours vs manual contours) is planned as an extension of this benchmark.
- **Deterministic single-model outputs.** Neither model outputs per-voxel uncertainty. Incorporating epistemic uncertainty via Monte-Carlo dropout, deep ensembles or evidential learning could support clinician review by flagging low-confidence regions, and would be particularly valuable given the small-primary failure mode identified here.

## Conclusions

On the HECKTOR 2025 Task 1 internal validation cohort, the choice between a 18 M-parameter custom 3D U-Net and an 88 M-parameter auto-configured nnU-Net v2 was worth 0.244 median GTVp DSC, and an adaptive post-processing stage recovered 79 % of that difference (0.192), leaving a residual three to four times smaller than the post-processing contribution itself. Without post-processing, nnU-Net v2 was decisively superior (median GTVp DSC 0.799 vs 0.592; d = –0.88); with it, the custom pipeline matched nnU-Net v2 on median GTVp DSC (0.800 vs 0.799) at ∼21 % of the parameters and 34 s versus 78 s per case, while remaining inferior on nodal disease after Bonferroni correction and failing catastrophically eight times more often on small primaries (8/98 vs 1/98; McNemar exact p = 0.016).

Two practical conclusions follow. For groups constrained to a lightweight custom network, inference-time post-processing recovers most of the accuracy that choice costs, and does so without retraining; for groups free to choose, the framework baseline remains the larger lever. For the field, pipeline-level comparisons reported without a post-processing ablation risk attributing to architecture an effect that belongs elsewhere; the ablation costs a fraction of the training it contextualises and should accompany such comparisons as standard. Neither conclusion supports deploying the lightweight pipeline without a review step targeting small primaries, where its failure mode is concentrated.

## Data Availability

All data are publicly available. The HECKTOR 2025 Task 1 FDG-PET/CT imaging data are distributed by the challenge organisers under the challenge's Data Use Agreement (https://hecktor25.grand-challenge.org/); no new patient data were collected by the authors. The complete analysis pipeline, both trained model checkpoints, the auto-configured nnU-Net plan, the fixed patient-level split, the completed CLAIM checklist, the internal adversarial review record and the verification suite that checks every reported figure against its generating script are archived at Zenodo, DOI 10.5281/zenodo.22147983.

https://doi.org/10.5281/zenodo.22147983

## Declarations

## Ethics approval and consent to participate

The authors declare that all relevant ethical guidelines have been followed. This study is a secondary computational analysis of publicly available, fully de-identified imaging data distributed by the HECKTOR 2025 MICCAI Challenge under the challenge’s Data Use Agreement; no new data were collected from human participants by the authors. Institutional review board approval and individual patient consent for the original acquisition were obtained by the contributing centres as a condition of the challenge’s data release, and are documented by the challenge organisers. No additional IRB or ethics committee approval was required for the present analysis, and no identifiable patient information was accessed at any point.

## Consent for publication

Not applicable.

## Availability of data and materials

The HECKTOR 2025 dataset is publicly available via the HECKTOR grand challenge portal (https://hecktor25.grand-challenge.org/) under the challenge’s Data Use Agreement.

Code is released in two parts. The **analysis pipeline** (nnU-Net v2 benchmarking, the three-arm statistical analysis with multiple-testing correction and McNemar tests, the table generators, per-patient CSV files, figure generation and a verification suite that checks every reported figure against its generating script) is parameterised and runnable, with pinned environment files. The **MiniUNet3D model, training loop and adaptive post-processing** are archived *verbatim* as extracted from the original research notebook, deliberately unrefactored so that the code which produced the published numbers remains auditable in the form it ran; a single personal filesystem path was redacted, and that is the only edit. Random seeds for split, weight initialisation and augmentation are included. All of it is deposited, together with the trained weights, on Zenodo with DOI **10.5281/zenodo.22147983**.

Trained model weights (MiniUNet3D and nnU-Net v2 checkpoints) are deposited on Zenodo with DOI **10.5281/zenodo.22147983**, in the same record as the analysis code. Random seed 42 was used throughout for reproducibility of the training and split; additional per-component seeds (data augmentation, TTA order) are documented in the code repository.

## Authors’ contributions

R.O.S. conceived the study, designed and implemented the MiniUNet3D architecture and post-processing pipeline, trained and benchmarked both models, performed the statistical analysis and drafted the manuscript. P.H.H. contributed to the implementation of the code and critically revised the manuscript. Both authors read and approved the final version.

## Use of artificial intelligence

In accordance with ICMJE recommendations, the authors disclose the use of a large-language-model assistant (Anthropic Claude, accessed through Claude Code) for assistance with programming and with drafting and revising the text of this manuscript. All model training, inference and statistical analysis were performed on hardware owned by the authors; no third-party computing service was used. The study was conceived, designed, executed and interpreted by the authors. Every design decision, every reported result and the final text are the authors’ own work and their sole responsibility. The assistant is not an author, and contributed no study design, no data and no scientific conclusion.

## Acknowledgements

The authors thank the HECKTOR challenge organisers for making the multicentric dataset available, and the nnU-Net authors for the open-source release of the framework.

## Reporting standards

This study is reported in accordance with the CLAIM (Checklist for AI in Medical Imaging) recommendations [26]; the completed checklist is Additional file 1, and items 29 (cohort demographics, unavailable in the public challenge release) and 39 (external validation) are declared as not met. Metrics were selected following the Metrics Reloaded framework [25].

